# Two and a Half Decades of Evidence on PTSD Determinants in Conflict Regions of Sub-Saharan Africa A Systematic Review and Meta-analysis

**DOI:** 10.64898/2026.02.28.26347310

**Authors:** Stewart Ngasa, Lionel Nges, Neh Ngasa, Therence Dingana, Sarmad Nadeem

**Author notes:** Corresponding author (SN).

## Abstract

Armed conflict in Sub-Saharan Africa has exposed millions of civilians to repeated and severe traumatic events, yet the prevalence of posttraumatic stress disorder (PTSD) and its associated determinants across the region have not been comprehensively synthesised. This study aimed to estimate the prevalence of PTSD and examine its associated factors among conflict-affected adult populations in Sub-Saharan Africa. Methodological quality was assessed using the Joanna Briggs Institute (JBI) criteria for cross-sectional and epidemiological studies A systematic search of PubMed, MEDLINE, Embase, Scopus, CINAHL, APA PsycINFO, the Cochrane Library, and the WHO Global Index Medicus (including African Index Medicus) was conducted for studies published between January 1, 2000, and May 31, 2025. Observational studies reporting PTSD prevalence among adults aged 18 years or older exposed to armed conflict were included. Study selection followed PRISMA 2020 guidelines, with independent screening by two reviewers. Random-effects meta-analyses with logit transformation were used to pool prevalence estimates, and determinants were synthesised narratively with emphasis on adjusted effect estimates. Heterogeneity was assessed using the I² statistic.

Sixty-eight studies comprising 82,021 participants from 13 countries met inclusion criteria. The pooled prevalence of PTSD was 43% (95% CI, 35.9%–50.0%), with substantial heterogeneity (I² = 99.9%). Prevalence was highest among refugees (79%), followed by internally displaced persons (48%) and residents of conflict-affected communities (34%). Female sex was consistently associated with increased odds of PTSD (pooled adjusted odds ratio approximately 2.0), as were comorbid depression or depressive symptoms (AOR range 4.2–9.5). Additional correlates included cumulative trauma exposure, displacement, poor social support, and substance use. Overall, PTSD is highly prevalent among conflict-affected adults in Sub-Saharan Africa, underscoring the need for integrated, context-sensitive mental health strategies to address the enduring psychological consequences of armed conflict in the region.

## Introduction

Armed conflict has affected dozens of countries across Sub-Saharan Africa over the past several decades, exposing millions of civilians to violence, displacement, and chronic insecurity. Globally, most people will experience at least one potentially traumatic event during their lifetime, yet only a minority will develop posttraumatic stress disorder (PTSD). Recent World Health Organization (WHO) estimates suggest that about 3.9% of the world’s population has experienced PTSD at some point in their lives, and approximately 5.6% of those exposed to trauma develop the disorder[1]. Among people exposed to war or other violent conflict, PTSD prevalence is substantially higher, reaching 15.3% in some estimates[1]. These figures indicate a pronounced concentration of PTSD in conflict-affected settings, where trauma exposure is widespread, recurrent, and often compounded by displacement and poverty.

Meta-analytic work over the last two decades has shown that conflict-affected populations carry a large and persistent burden of mental disorders, including PTSD. A landmark systematic review of 181 surveys among more than 81 000 refugees and conflict-exposed civilians across 40 countries reported an unadjusted weighted PTSD prevalence of 30.6% (95% CI, 26.3%–35.2%)[2]. More recent WHO-supported modelling of conflict settings estimated that 22.1% of adults in these contexts live with a mental disorder at any point in time, including depression, anxiety, PTSD, bipolar disorder, and schizophrenia[3]. A global epidemiological model further suggests that 316–354 million adult war survivors worldwide are living with PTSD and/or major depression[3–5]. Within war-affected civilian populations, pooled PTSD prevalence estimates in recent reviews typically range from approximately 23% to more than 30%, again with marked heterogeneity across settings[3,5].

In Sub-Saharan Africa, recurrent and protracted conflicts in countries such as Ethiopia, South Sudan, Democratic Republic of Congo, Nigeria, Cameroon, Uganda, Rwanda, and Mozambique have created large populations exposed to extreme and chronic violence[6–11]. A growing number of epidemiologic and clinical studies from the region—including those conducted among internally displaced persons, refugees, ex-combatants, survivors of sexual and gender-based violence, and conflict-affected community samples—have documented high PTSD symptom burden and frequent comorbid depression and anxiety [12–20]. Many of these studies report PTSD prevalence estimates substantially higher than global community averages and consistently identify risk factors such as cumulative trauma exposure, forced displacement, low social support, and poverty [12–20].

Beyond estimating prevalence, numerous studies have examined factors associated with PTSD in conflict-affected African populations. Frequently reported correlates include female sex, comorbid depression, exposure severity, intimate-partner violence, ongoing insecurity, and barriers to mental health care. However, findings are dispersed across many cross-sectional studies with heterogeneous methodologies, which makes it difficult to determine the relative strength of associations across the region.

Despite more than two decades of research, there is no comprehensive synthesis quantifying PTSD prevalence and associated factors across the full breadth of conflict-affected countries in Sub-Saharan Africa. Existing global or regional reviews often aggregate data across dissimilar settings, potentially obscuring region-specific patterns. By focusing on Sub-Saharan Africa exclusively, the present review aims to provide a more contextually grounded understanding of PTSD epidemiology in conflict settings.

Therefore, the objective of this systematic review and meta-analysis was to synthesize two and a half decades (2000–2025) of evidence on PTSD among conflict-affected adults in Sub-Saharan Africa. Specifically, we aimed to (1) estimate pooled PTSD prevalence across the region and (2) quantify the associations between PTSD and reported determinants. Given that most primary studies are cross-sectional, we use the term “determinants” to denote factors statistically associated with PTSD, acknowledging that these should be interpreted as associated factors rather than proven causal agents. By integrating findings across multiple countries and conflict contexts, this review provides the most extensive assessment to date of the burden and correlates of PTSD in Sub-Saharan Africa.

## Materials and Methods

### Protocol and reporting

This systematic review and meta-analysis were conducted and reported in accordance with the PRISMA 2020 statement and the MOOSE (Meta-analysis of Observational Studies in Epidemiology) guidelines for meta-analyses of observational studies[21]. The protocol was prospectively registered in the International Prospective Register of Systematic Reviews (PROSPERO; registration number CRD420251020426) before the review commenced. All subsequent methodological decisions were guided by this protocol, and any minor refinements were consistent with the original objectives.

### Eligibility criteria and research question

The review addressed the question: “What is the prevalence of post-traumatic stress disorder (PTSD) and its determinants among conflict-affected adult populations in Sub-Saharan Africa?” Studies were eligible if they involved adults aged 18 years or older who had experienced war or armed conflict and were living in conflict-affected settings within countries classified as Sub-Saharan Africa. Eligible populations included internally displaced persons (IDPs), refugees, and residents or survivors living in communities affected by conflict. Only observational cross-sectional studies reporting data on the prevalence of PTSD were included in the meta-analysis, and PTSD had to be assessed using a clearly defined diagnostic or screening instrument with an explicit diagnostic algorithm or cut-off score. Studies that also reported determinants of PTSD were included in the narrative synthesis of risk factors if they provided quantitative measures of association. To ensure the relevance and comparability of the assembled evidence, inclusion was restricted to primary quantitative studies conducted in Sub-Saharan African countries, published in peer-reviewed journals in English between 1 January 2000 and the date of the final search. Studies were excluded if they lacked sufficient data to calculate PTSD prevalence or extract effect estimates, were not primary quantitative research, or focused exclusively on participants younger than 18 years.

### Information sources and search strategy

A comprehensive search strategy was implemented to identify relevant observational studies. Two independent researchers, in collaboration with an experienced information specialist, developed and conducted the searches. The following electronic databases were searched: PubMed, MEDLINE, CINAHL, the Cochrane Library, Embase, APA PsycINFO, and the WHO Global Index Medicus (including the African Index Medicus). Searches covered publications from 1 January 2000 up to the date of the final search. Search strategies were tailored to each database and combined controlled vocabulary terms (such as MeSH in PubMed and Emtree in Embase) with free-text keywords. Search terms reflected three main conceptual domains: PTSD and related mental health constructs; exposure to war, armed conflict, and forced displacement; and epidemiological outcomes, including prevalence and associated factors. Illustrative terms for PTSD included “post-traumatic stress disorder”, “PTSD”, and “stress disorder, post-traumatic”. Terms capturing conflict and displacement included “war”, “armed conflict”, “war zone”, “civil war”, “genocide”, “refugee”, “internally displaced persons”, and “displaced”. Epidemiological terms included “prevalence”, “incidence”, “risk factors”, “determinants”, and “epidemiology”.

### Study selection

All records identified from the database searches and reference list screening were imported into Microsoft Excel and de-duplicated. Study selection was conducted in two stages. In the first stage, two reviewers independently screened titles and abstracts against the predefined eligibility criteria, excluding records that clearly did not meet the inclusion criteria. In the second stage, the full texts of all potentially relevant articles were obtained and independently assessed for eligibility by the same two reviewers. Disagreements at any stage were resolved through discussion; if consensus could not be reached, a third senior reviewer was consulted. Reasons for exclusion at the full-text stage were documented systematically to provide a transparent account of the selection process. The final set of included studies was agreed by the full review team, ensuring that the assembled body of evidence was appropriate to address the review question in line with MOOSE recommendations.

### Data extraction

Data were extracted using a customised extraction form designed in Microsoft Excel. The form was developed based on the objectives of the review and piloted on a subset of included studies to ensure clarity and consistency before being applied to all articles. Two reviewers independently extracted data from each study. Extracted variables included study identification (first author, year of publication), country and setting, study design, sampling strategy where reported, and characteristics of the study population (sample size, age distribution when available, and classification of the population as internally displaced persons, refugees, or residents/survivors). Information on the PTSD assessment instrument used (including version and language where reported), the operational definition or cut-off used to define PTSD, and the reported prevalence of PTSD overall and by relevant subgroups (for example, by sex or population group) was also extracted. For studies reporting determinants of PTSD, data were collected on potential risk and protective factors, including socio-demographic characteristics, conflict-related exposures, psychosocial and contextual variables, and mental health comorbidities, together with corresponding effect estimates and 95% confidence intervals. Where multiple statistical models were presented, adjusted effect estimates were preferentially extracted, and the covariates included in each model were recorded to allow explicit consideration of confounding in the interpretation of findings. Discrepancies between reviewers in extracted data were resolved by returning to the original articles and discussing the differences within the review team. When essential data were missing or unclear, attempts were made to contact study authors for clarification or additional information; if no response was obtained after reasonable efforts, missing data were documented, and the study was retained with appropriate notation of limitations in the synthesis.

### Assessment of methodological quality

The methodological quality and risk of bias of the included studies were appraised using the Joanna Briggs Institute tool for epidemiological proportional, non-comparative studies[22]. This tool addresses biases in sample frame adequacy, sampling methodology, sample size justification, validity and reliability of measurement tools, case definition clarity, standardization of measurements, response rate, and handling of missing data. Each of the nine questions was answered with “yes”, “no”, or “unclear”. Each “yes” response scored 1 point, while “no” or “unclear” responses scored 0, yielding a total score between 0 and 9. Four reviewers participated in the quality assessment process, with each study independently assessed by at least two reviewers’ familiar with epidemiological methods and mental health outcomes. Studies were categorized a priori as low quality (scores ≤4), moderate quality (scores 5–6), or high quality (scores 7–9). Disagreements in quality ratings were resolved through discussion and, when necessary, consultation with another reviewer until consensus was reached. Particular attention was paid to the comparability domain, which reflects the extent to which analyses accounted for important confounding variables such as age, sex, and indicators of conflict exposure. Study quality ratings were not used as exclusion criteria but were incorporated into the interpretation of findings, including sensitivity analyses and narrative discussion of the robustness and consistency of evidence.

### Data synthesis and statistical analysis

Quantitative synthesis was undertaken for studies that reported sufficient data to estimate PTSD prevalence in the target populations. The primary effect measure was the prevalence of PTSD, defined as the proportion of participants meeting the diagnostic criteria or cut-off specified for the PTSD instrument used. When necessary, prevalence estimates and standard errors were calculated from the reported numbers of cases and total sample sizes. Because methodological and contextual heterogeneity across studies was anticipated, pooled prevalence estimates and corresponding 95% confidence intervals were calculated using random-effects meta-analysis with logit transformation of proportions for between-study variance. Between-study heterogeneity was quantified using τ² and Higgins’ I² statistic, and 95% prediction intervals were estimated to describe the expected range of true PTSD prevalence across similar populations beyond those included in the meta-analysis.

Where data permitted, subgroup analyses were conducted by country and by affected group, distinguishing between internally displaced persons, refugees, and residents or survivors living in conflict-affected communities, to explore potential sources of heterogeneity. Determinants of PTSD were synthesized narratively, with factors grouped into conceptual categories such as sociodemographic characteristics, conflict-related exposures, and psychosocial variables. Emphasis was placed on adjusted effect estimates where available. Owing to variability in the measurement and reporting of determinants and in the covariates included in multivariable models, and the resulting limited comparability of effect estimates across studies, formal meta-regression was not undertaken. Potential small-study effects and publication bias were examined, where at least ten studies were available in a meta-analysis, using visual inspection of funnel plots and Egger’s regression test. All statistical analyses were performed using Jamovi statistical software version 2.644,[23], (Sydney, Australia), and key analytical decisions were checked by at least two independent reviewers to ensure accuracy and adherence to the prespecified analysis plan.

## Results

### Search strategy

A total of 4,671 records were identified through database searching. After the removal of 1,318 duplicates, 3,301 unique titles and abstracts were screened. Of these, 103 full-text articles were retrieved and assessed for eligibility, resulting in 68 studies being included in the review and meta-analysis. The main reasons for exclusion at the full-text stage were inaccessible full text, studies being errata or protocols, discordance between the intervention or population and the review question, lack of reported PTSD prevalence data, and duplicate publication of the same data, as shown in the PRISMA flow diagram (Figure 1).

**Figure 1.**
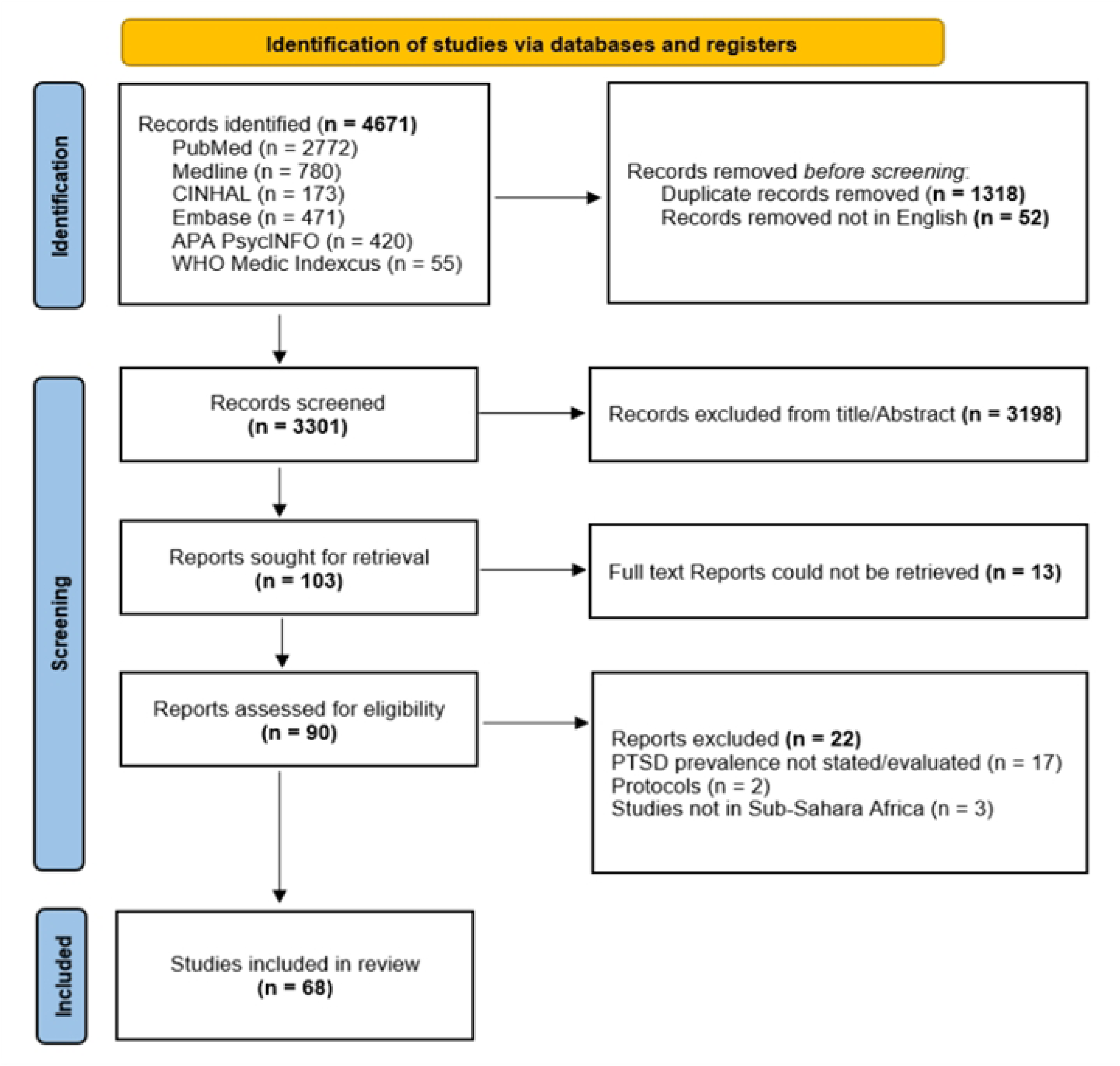
PRISMA Flow diagram

### Characteristics of included studies

The 68 included studies comprised a combined sample of 82,021 adult participants from conflict-affected settings in Sub-Saharan Africa. Of these, 30,129 (36.7%) were male, of the total sample. Studies were conducted in Ethiopia (15),[6,20,24–36] Uganda (13), [13,17,37–47] Sudan (9),[48–56] Rwanda (8),[9,57–63] Nigeria (6),[8,18,64–67] the Democratic Republic of Congo, DRC (6),[11,15,16,19,68,69] Somalia (3),[26,70,71] South Sudan (2),[7,72] Kenya (2),[73,74] Liberia (2),[12,14] Côte d’Ivoire (1),[75] and Mozambique (1)[10] (Figure 2).

**Figure 2.**
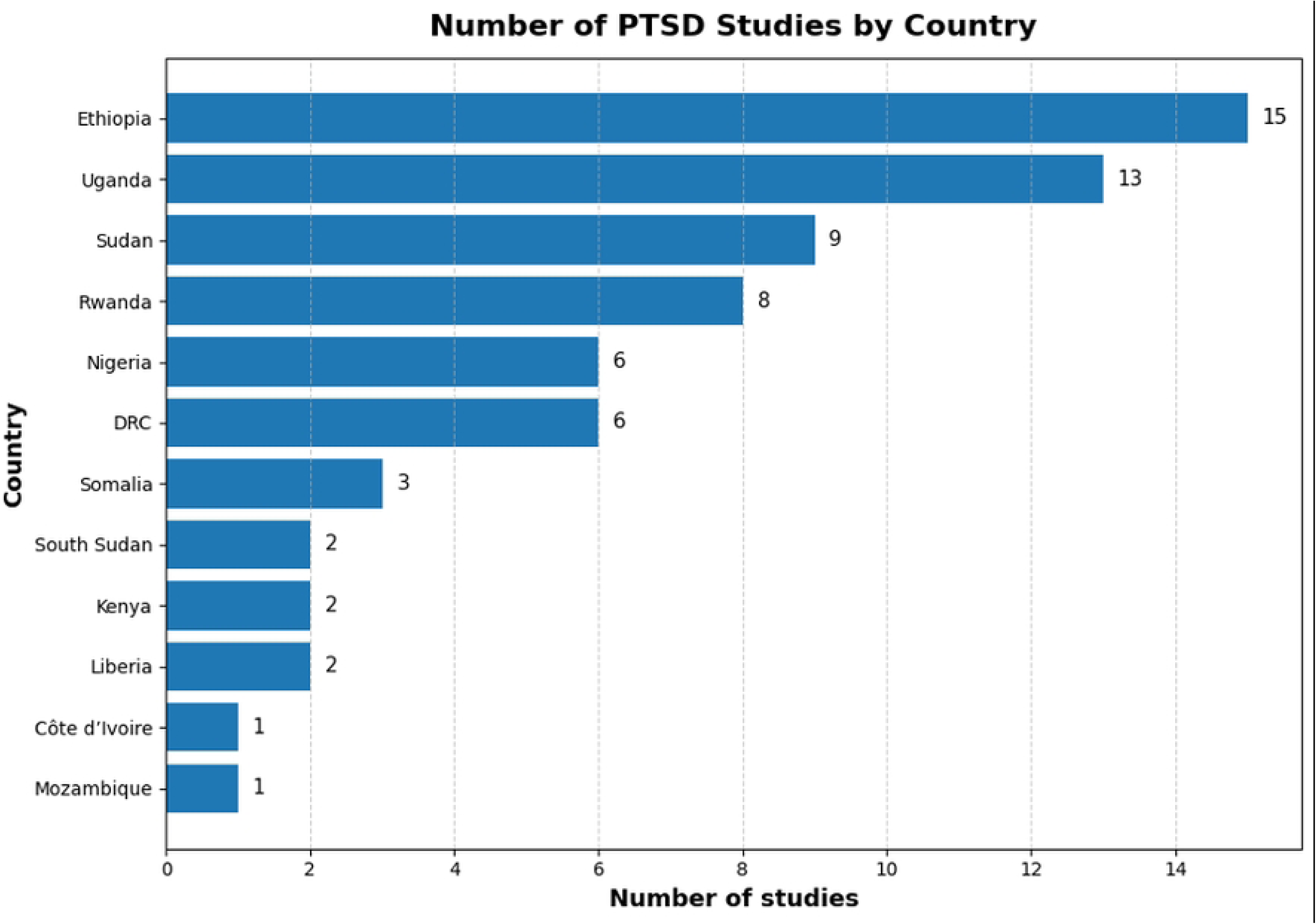
Study distribution across countries in the sub-Sahara.

The included studies were published between 2000 and the date of the final search and were conducted in various conflict-related settings, including internally displaced persons (IDP) camps, refugee camps or settlements, and communities affected by conflict. Regarding population focus, 16 studies were conducted among IDPs, 10 among refugees, and 42 among residents or survivors living in conflict-affected communities. PTSD was assessed using a range of diagnostic and screening instruments, most commonly the PTSD Checklist (PCL-5, 15 studies; PCL-C, 8 studies) and the Harvard Trauma Questionnaire (HTQ, 13 studies). Other tools included the Mini International Neuropsychiatric Interview (MINI, 8 studies), the Posttraumatic Diagnostic Scale (PDS, 5 studies), the Impact of Event Scale (IES, 3 studies), the Composite International Diagnostic Interview (CIDI, 2 studies), and the PTSD Symptom Scale (PSS, 2 studies). Some studies did not clearly report the instrument used.

### Risk Assessment

Methodological quality was assessed using the Joanna Briggs Institute (JBI) criteria for cross-sectional and epidemiological studies, with a maximum score of 9 points [22]. This was an overall high-quality review, as most studies had a low risk of bias (44 studies), moderate risk (19 studies), and high risk (5 studies) (supplementary sheet 1).

### Socio-demographic Characteristics of participants

Across studies, participants included conflict-affected civilians, comprising internally displaced persons (IDPs), refugees, and non-displaced residents living in areas affected by armed conflict. Several studies focused exclusively on IDPs, whereas others enrolled mixed populations that included displaced and nondisplaced individuals. Sample sizes ranged from approximately 364 to more than 1,000 participants. Participant age was reported in most studies, commonly as a mean with SD or as a median with interquartile range. Where reported, mean ages generally fell within early to mid-adulthood. Sex distribution was reported in the majority of studies. The proportion of female participants varied widely across samples, ranging from approximately 18% to 47%, with males comprising the remaining proportion.

Reported prevalence estimates of the primary outcome varied substantially across studies and populations. Prevalence values ranged from low to high proportions, with several studies reporting estimates exceeding 30%, particularly among IDPs and populations exposed to prolonged conflict or displacement. Lower prevalence estimates were observed in studies including mixed or nondisplaced populations (Supplementary sheet 2).

### PTSD outcome

Across all 68 studies, the pooled prevalence of PTSD in conflict-affected adult populations in Sub-Saharan Africa was 0.43 (95% CI 0.38–0.49), corresponding to an overall prevalence of 43%. Statistical heterogeneity between studies was very high, with an I² of 99.5% and a between-study variance (τ²) of 0.06, and a Prediction interval PI (0.06 - 0.87). Table 1 and Figure 3

**Figure 3:**
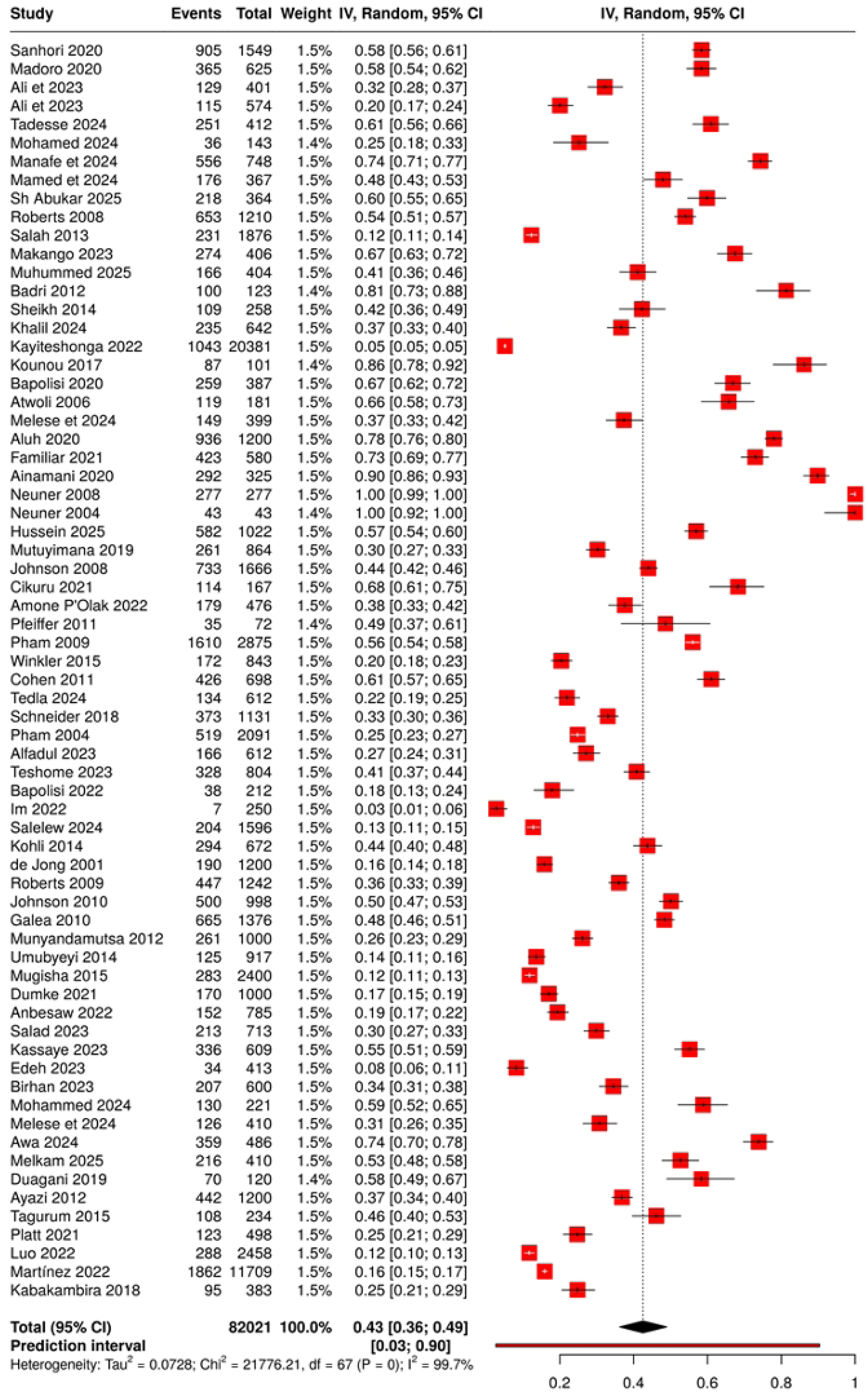
Forest plot of all 68 included studies.

**Table 1:** Pooled PTSD prevalence by country.

| Countries | Studies | Prevalence | 95%CI LL | 95%CI UL |
| --- | --- | --- | --- | --- |
| Ethiopia | 15 | 0.397 | 0.303 | 0.492 |
| Uganda | 13 | 0.54 | 0.309 | 0.772 |
| Sudan | 9 | 0.418 | 0.262 | 0.574 |
| Rwanda | 8 | 0.263 | 0.145 | 0.38 |
| Nigeria | 6 | 0.461 | 0.419 | 0.504 |
| DRC | 6 | 0.424 | 0.261 | 0.587 |
| Somalia | 3 | 0.406 | 0.23 | 0.581 |
| South Sudan | 2 | 0.364 | 0.345 | 0.383 |
| Kenya | 2 | 0.342 | -0.275 | 0.959 |
| Liberia | 2 | 0.461 | 0.419 | 0.504 |
| Côte d'Ivoire | 1 | 0.861 |  |  |
| Mozambique | 1 | 0.743 | 0.7104 | 0.7742 |
| **Pooled Prevalence** | **68** | **0.429** | **0.359** | **0.5** |

Note. CI; Confidence intervals, LL; lower limit, UL; upper limit.

### Sub-group Analysis by Country

A subgroup analysis was conducted by country, and heterogeneity within each subgroup remained high, with I² ranging from 82.49% to 99.94%, (Table 2). Country-specific results are presented in detail using a geographical map to illustrate the distribution of PTSD prevalence across Sub-Saharan Africa and shows a spatial distribution with higher prevalences in the eastern regions of sub-Saharan Africa (Figure 4). Generally, countries with multiple studies, such as Ethiopia, Uganda, Sudan, Rwanda, Nigeria, and the Democratic Republic of Congo, demonstrated consistently high levels of PTSD, although point estimates varied substantially between individual studies, even within the same country. Subgrouping by country did not meaningfully reduce heterogeneity, again highlighting the influence of differences in study design, measurement instruments, and population characteristics.

**Figure 4:**
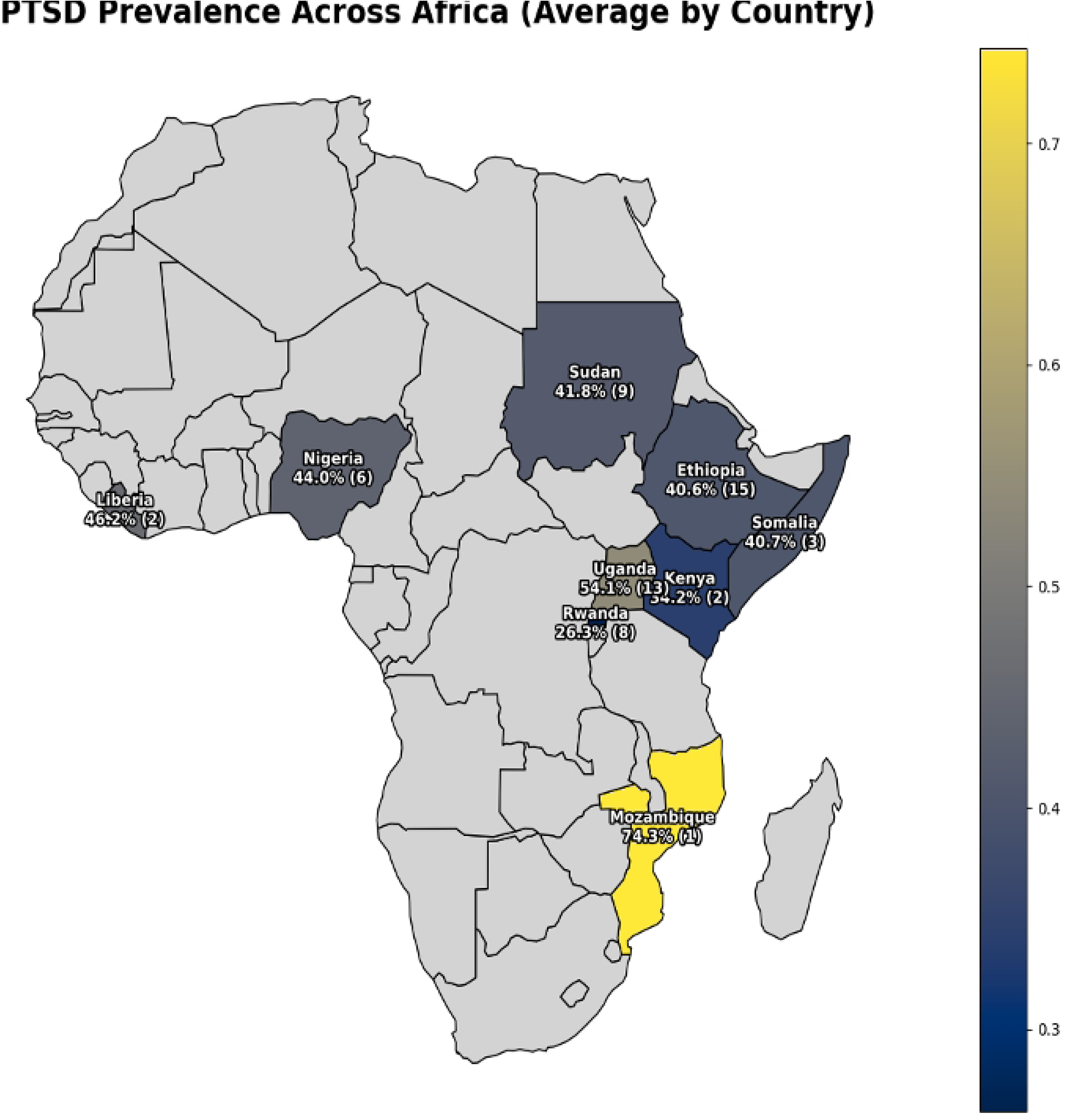
Geo-spatial representation of PTSD prevalence across different countries in sub-Saharan Africa

**Table 2:**
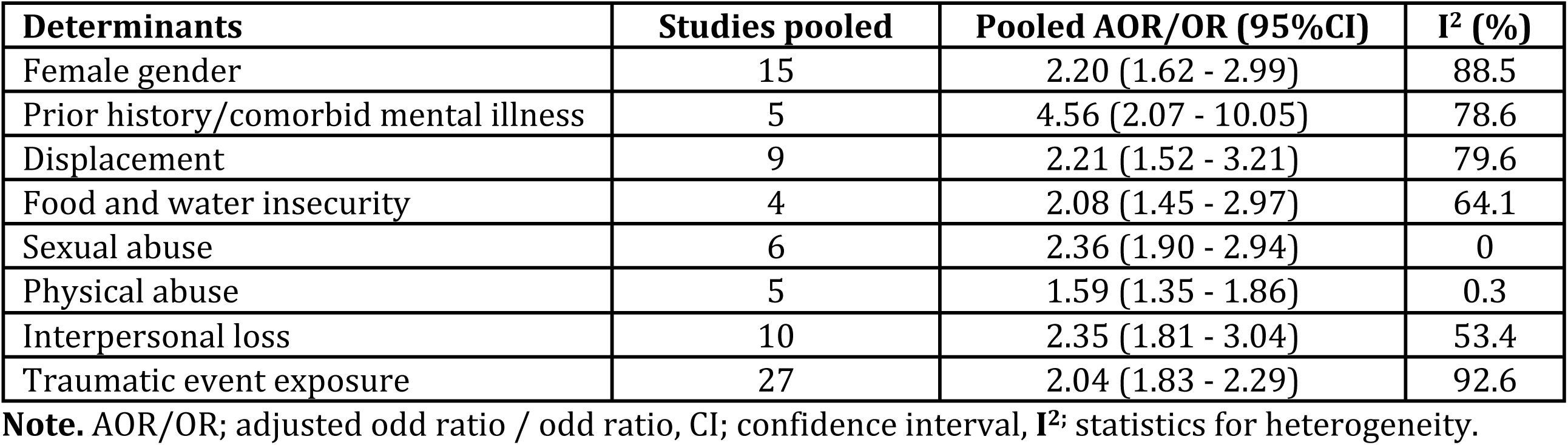
Pooled results of selected PTSD determinants.

| Determinants | Studies pooled | Pooled AOR/OR (95%CI) | $I^2$ (%) |
| --- | --- | --- | --- |
| Female gender | 15 | 2.20 (1.62 - 2.99) | 88.5 |
| Prior history/comorbid mental illness | 5 | 4.56 (2.07 - 10.05) | 78.6 |
| Displacement | 9 | 2.21 (1.52 - 3.21) | 79.6 |
| Food and water insecurity | 4 | 2.08 (1.45 - 2.97) | 64.1 |
| Sexual abuse | 6 | 2.36 (1.90 - 2.94) | 0 |
| Physical abuse | 5 | 1.59 (1.35 - 1.86) | 0.3 |
| Interpersonal loss | 10 | 2.35 (1.81 - 3.04) | 53.4 |
| Traumatic event exposure | 27 | 2.04 (1.83 - 2.29) | 92.6 |

**Note.** AOR/OR; adjusted odd ratio / odd ratio, CI; confidence interval, $I^2$ ; statistics for heterogeneity.

### Subgroup analysis by population

Subgroup analyses by population group revealed significant differences in estimated prevalence. Sixteen studies reported PTSD among IDPs, with a pooled prevalence 0.48 (95% CI 0.37–0.59; I² = 99.3% (95%CI; 98.4% – 99.7%); τ² = 0.05 (95%CI; 0.02–0.09). Among refugees (10 studies), the pooled prevalence was higher, at 0.74 (95%CI; 0.38 – 0.97), I² = 99.9% (95%CI; 99.8% – 99.9%; τ² ≈ 0.33). In contrast, residents and survivors living in conflict-affected communities (42 studies) had a lower, though still substantial, pooled PTSD prevalence of 0.33 (95%CI; 0.28–0.39, I² = 99.3%, 95%CI; 99.3% – 99.7%, τ² = 0.03, 95% CI 0.02–0.05). Overall, these subgroup analyses suggested a gradient, with refugees showing the highest PTSD prevalence, followed by IDPs, and then residents or survivors. However, the degree of heterogeneity within each subgroup remained similarly high and did not decrease compared with the overall analysis.

### PTSD determinants

Across the 68 included studies, approximately 50 distinct determinants of PTSD were evaluated. Of these, 55 studies examined determinants, while 37 conducted quantitative analyses reporting adjusted odds ratios or odds ratio estimates (AOR/OR). The determinants evaluated in these 37 studies were grouped into three broad domains: individual-level, socioeconomic, and trauma-related factors. Individual-level determinants included age, marital status, educational attainment, religion or religious affiliation, and gender. Female sex emerged as a near-universal determinant of PTSD across settings and populations. Socioeconomic determinants encompassed employment status, income level or wealth index, food insecurity, and displacement-related factors, including refugee or internally displaced person (IDP) status, duration of displacement, and living conditions within displacement camps. Trauma-related determinants included cumulative trauma exposure, witnessing killings or severe violence, direct combat exposure, kidnapping or abduction, and severe human rights violations such as sexual violence or rape, and physical abuse or torture. Trauma-related factors consistently showed the strongest associations with PTSD, with reported effect sizes frequently exceeding an AOR/OR of 3.0.

Pooled determinants of PTSD are summarised in Table 2 and shown in Figures 6 and 7. Eight determinants were evaluated across the included studies: female gender, history or comorbid mental illness, displacement, food insecurity, sexual abuse or violence, physical abuse, violence or torture, interpersonal loss, and overall trauma exposure. These factors were selected because they were the most consistently reported across studies and provided quantifiable data suitable for pooled analysis. Other potential determinants were not included due to limited reporting or heterogeneous measurement, which would have compromised the reliability of pooled estimates.

**Figure 5.**
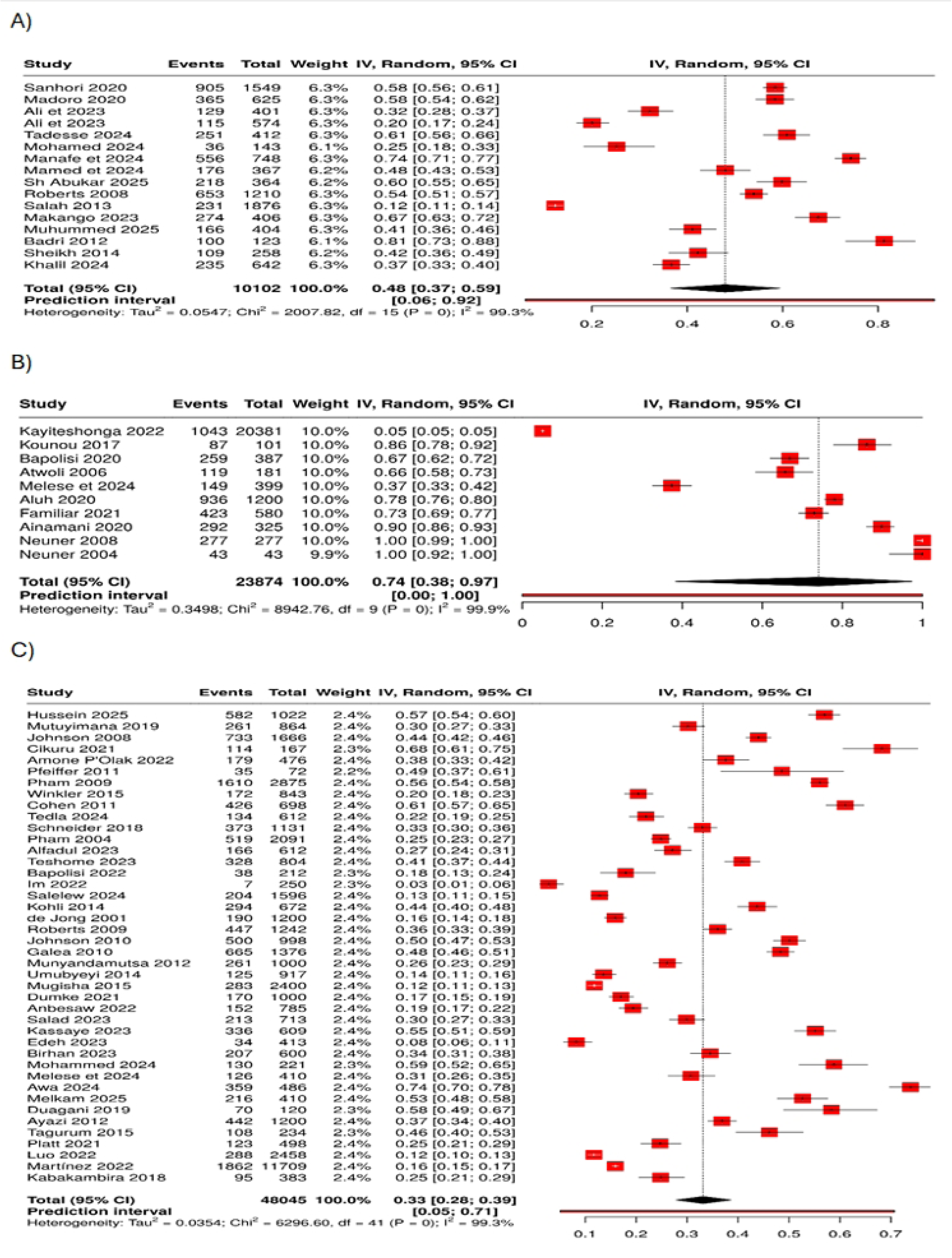
Forest plot showing subgroup analyses by different population groups. A) IDP forest plot, B) Refugee Forest plot, C) residents/survivors Forest plot.

**Figure 6.**
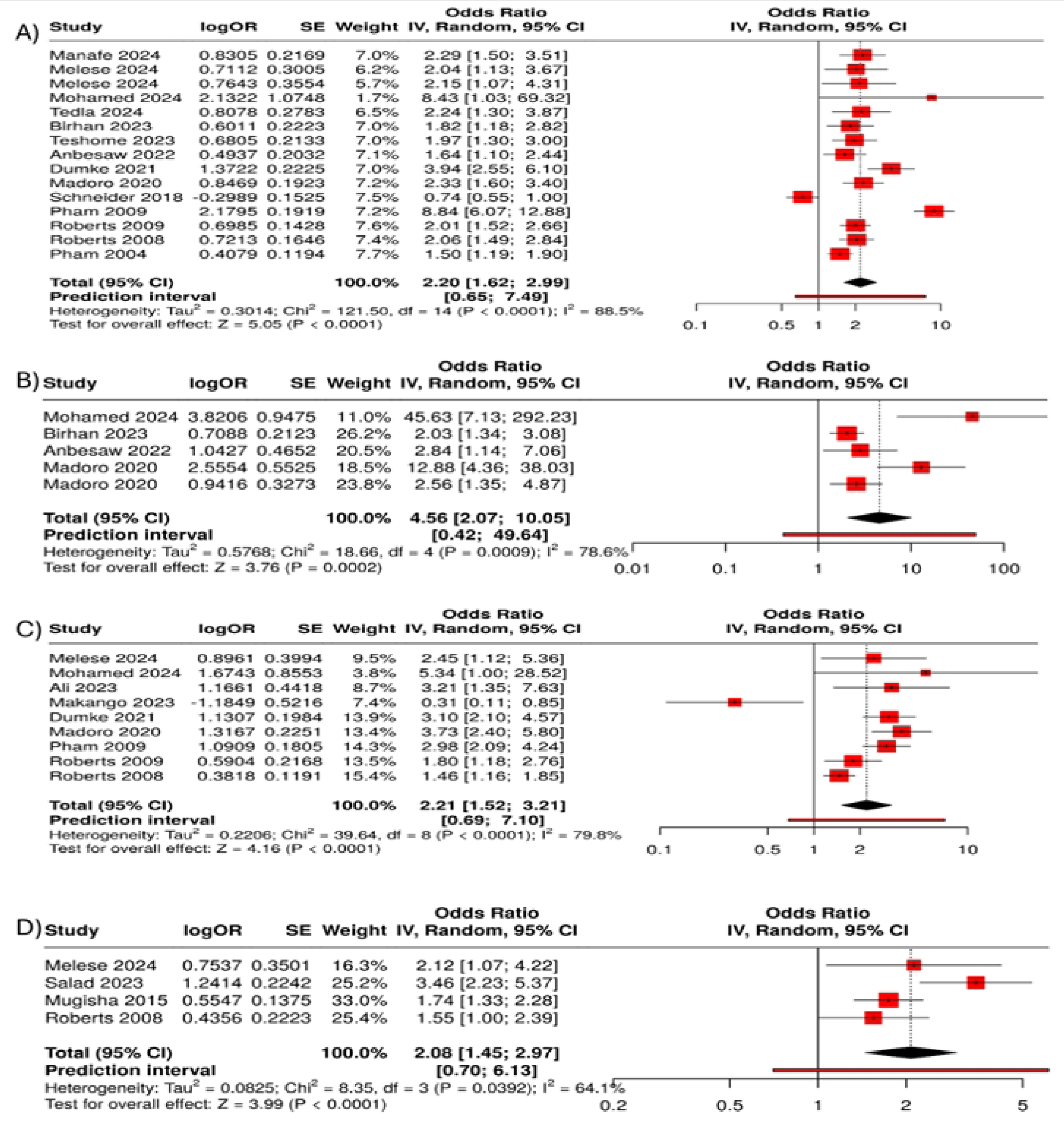
Forest plots of individual level and socioeconomic determinants of ptsd. A) female gender, B) prior history of or comorbid mental illness, C) displacement, D) Food insecurity.

**Figure 7:**
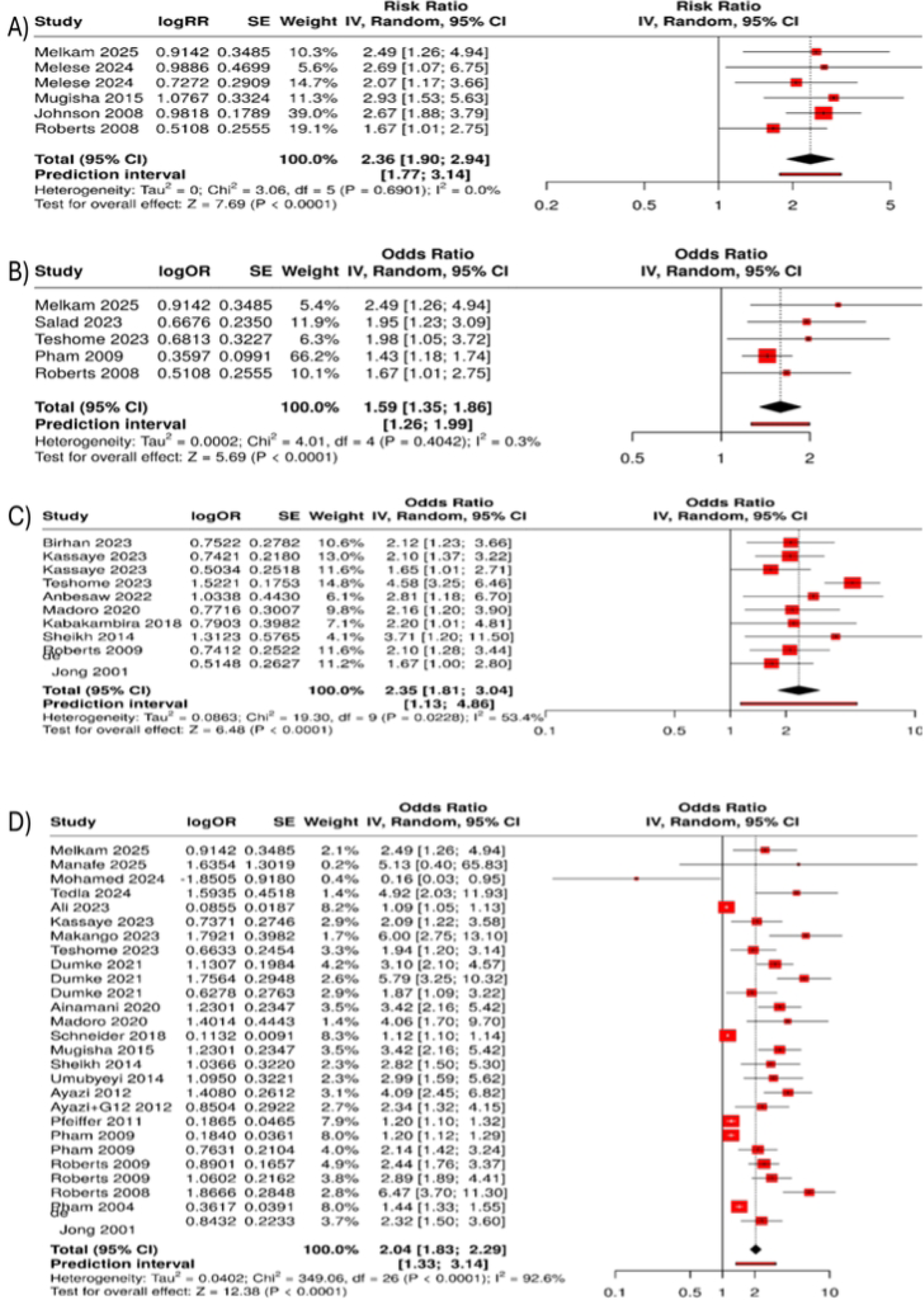
Forest plots showing trauma related determinants of PTSD. A) sexual violence/abuse/rape, B) physical violence/abuse/torture, C) interpersonal loss or witness of loss of family/loved one or friend, D) trauma exposure.

## Discussion

In this systematic review and meta-analysis of 68 observational studies conducted over a 25-year period, we found that posttraumatic stress disorder (PTSD) is highly prevalent among conflict-affected adult populations in Sub-Saharan Africa, with a pooled prevalence of 43%. This estimate substantially exceeds global lifetime prevalence estimates for PTSD of 3.9%[76] and is higher than most pooled estimates reported in conflict-affected populations worldwide[[3]. Together, these findings indicate a substantial and persistent burden of trauma-related psychopathology in the region.

### Comparison With Prior Evidence

The prevalence observed in this review is notably higher than pooled estimates reported in earlier global meta-analyses of war-affected populations, which typically range from 15% to 30%[3]. WHO-supported epidemiological modelling studies have further estimated that approximately one in five adults in conflict settings experiences a mental disorder at any point in time, including PTSD, depression, and anxiety[1]. The higher prevalence identified in Sub-Saharan Africa may reflect the protracted and recurrent nature of conflicts in the region, as well as the high frequency of civilian exposure to violence, displacement, and chronic insecurity. Unlike conflicts characterised by discrete periods of violence followed by relative stability, many conflicts in Sub-Saharan Africa involve prolonged exposure to trauma and ongoing stressors, which are known to increase both PTSD risk and symptom persistence. [3,11,45,49,53].

### Heterogeneity and Measurement Considerations

As expected, between-study heterogeneity was substantial (I² >99%) and remained high across country-level and population subgroup analyses. This degree of heterogeneity is consistent with prior meta-analyses of PTSD prevalence in humanitarian settings and reflects considerable variation in study populations, conflict contexts, timing of assessment, and measurement approaches[2,5]. PTSD was assessed using a wide range of instruments, including screening questionnaires and structured diagnostic interviews, often with differing cut-off scores and variable cultural adaptation. Screening instruments may capture broader psychological distress related to ongoing adversity, potentially inflating prevalence estimates compared with diagnostic interviews.

Contextual factors also likely contributed to heterogeneity. Even within the same country, studies differed with respect to conflict intensity, displacement patterns, living conditions, and access to services. In countries like Sudan, Somalia and Ethiopia which have been plagued by several conflicts over the years. These conflicts occur for different reasons and may contribute to the high level of heterogeneity. The persistence of high heterogeneity despite subgroup analyses suggests that PTSD prevalence in conflict-affected Sub-Saharan Africa is highly context-specific and shaped by complex interactions between trauma exposure, post-conflict environments, and individual vulnerability. Accordingly, pooled prevalence estimates should be interpreted as indicators of overall burden rather than precise estimates applicable to all settings.

### Population Differences and Displacement

Subgroup analyses demonstrated a gradient in PTSD prevalence by population group, with refugees exhibiting the highest pooled prevalence, followed by internally displaced persons (IDPs), and then residents or survivors living in conflict-affected communities. This pattern is consistent with models conceptualising displacement as a cumulative stressor that compounds prior trauma exposure with additional adversities, including loss of social networks, legal insecurity, and prolonged uncertainty as demonstrated in previous reviews [1,2]. Refugees often experience extended displacement and limited access to resources, which may contribute to sustained PTSD symptoms [17,28,45,53,74,75].

However, the substantial prevalence observed among residents and survivors who remain in conflict-affected communities underscores that displacement alone does not account for the burden of PTSD. Ongoing exposure to insecurity, poverty, and structural deprivation may sustain psychological distress even among non-displaced populations [53]. These findings suggest that mental health needs extend beyond refugee and IDP populations and should be addressed within conflict-affected communities more broadly.

### Determinants of PTSD

The synthesis of determinants highlights PTSD in conflict-affected Sub-Saharan Africa as a condition influenced by both individual-level and contextual factors. Female sex emerged as one of the most consistent correlates, with women generally exhibiting approximately two-fold higher odds of PTSD compared with men. This finding aligns with global evidence and may reflect gendered patterns of trauma exposure, including higher rates of sexual and gender-based violence, as well as social and economic inequalities that may limit recovery opportunities [1,2]. Comorbid depression and depressive symptoms were among the strongest correlates of PTSD across studies, often demonstrating large effect sizes. This association likely reflects shared neurobiological mechanisms, overlapping symptomatology, and reciprocal reinforcement between PTSD and depression [77]. In contexts of chronic adversity, depressive symptoms may exacerbate trauma-related distress and functional impairment, contributing to persistent morbidity.

Trauma-specific exposures—including sexual violence, torture, physical assault, and abduction—were consistently associated with increased PTSD risk, with several studies demonstrating dose–response relationships between cumulative trauma exposure and PTSD [2]. Importantly, this review also highlights the independent contribution of non-violent conflict-related adversities, such as food insecurity, loss of housing, and property destruction. These findings support broader conceptualisations of PTSD in humanitarian settings that emphasise the role of sustained stressors and material deprivation alongside discrete traumatic events [1].

### Psychosocial and Behavioural Correlates

Psychosocial factors were prominent determinants of PTSD risk. Poor social support, disrupted family structures, and social isolation were consistently associated with higher odds of PTSD, while stable family and community relationships appeared protective. In many Sub-Saharan African contexts, where social connectedness plays a central role in coping and recovery, conflict-related disruption of social networks may have particularly adverse psychological consequences. Substance use, including alcohol and khat use, was also frequently associated with PTSD. These associations likely reflect bidirectional relationships, whereby individuals use substances to cope with trauma-related symptoms, while substance use may in turn exacerbate PTSD severity and impair recovery. These findings highlight the need to consider co-occurring substance use in clinical and public health responses to PTSD in conflict-affected populations.

### Policy and Practice Implications

The findings of this review have implications for mental health service organisation in low-resource, conflict-affected settings. The high prevalence of PTSD and its strong comorbidity with depression support the prioritisation of trauma-related disorders within primary care and community-based services. Consistent with the World Health Organization’s Mental Health Gap Action Programme (mhGAP)[78] task-sharing approaches—where trained non-specialist health workers deliver structured psychological interventions under supervision—may represent a feasible strategy to reduce the treatment gap. Stepped-care models integrating identification, brief psychosocial interventions, and referral pathways for more severe cases may be particularly relevant in settings with limited specialist capacity.

### Limitations

This review has several strengths, including its large cumulative sample size, exclusive focus on Sub-Saharan Africa, and rigorous adherence to PRISMA and MOOSE guidelines. By synthesising both prevalence and determinants, it provides a comprehensive assessment of PTSD burden and associated factors across diverse conflict settings.

However, most included studies were cross-sectional, limiting causal inference. Measurement of heterogeneity and reliance on screening instruments may have contributed to variability in prevalence estimates. Some conflict-affected countries remain underrepresented, and publication bias cannot be excluded. Despite these limitations, the consistency of key associations across heterogeneous settings supports the robustness of the principal findings.

### Conclusions

In summary, this systematic review and meta-analysis demonstrates that PTSD is highly prevalent among conflict-affected adults in Sub-Saharan Africa, with prevalence estimates substantially exceeding global averages. PTSD is consistently associated with female sex, comorbid depression, cumulative trauma exposure, displacement, and poor social support. These findings underscore the need for integrated, context-sensitive mental health strategies to address the long-term psychological consequences of armed conflict in the region

## Data Availability

The data underlying the results presented in the study are available from the corresponding author, Stewart Ngasa on the following

## Acknowledgment

Drs Stewart Ngasa and Nges had full access to all the data in the study and take responsibility for the integrity of the data and the accuracy of the data analysis. Drs Stewart Ngasa and Nges contributed equally to the conception and design of the study, independently screened and selected studies, conducted the data analysis, and drafted the manuscript. Drs Neh Ngasa, Dingana, and Nadeem contributed to study conception, critically reviewed the manuscript for important intellectual content, and approved the final version for publication. All authors read and approved the final manuscript.

## Additional Contributions

We thank the staff of the Pennine Care NHS Foundation Trust Library, particularly Stephen Edwards and Matt Johnson, for their assistance in identifying and retrieving full-text articles that were not accessible through online databases. We are especially grateful to Stephen Edwards for his time in discussing the review methodology and agreeing on a plan for completion of the project. No compensation was received for these contributions.

